# Hospital Perinatal Transmission Dynamics of Antimicrobial Resistance, Bangladesh, 2020

**DOI:** 10.1101/2022.06.28.22276992

**Authors:** Ashley Styczynski, Mohammed Badrul Amin, Shahana Parveen, Abu Pervez, Dilruba Zeba, Akhi Akhter, Helen Pitchik, Mohammad Aminul Islam, Muhammed Iqbal Hossain, Sumita Rani Saha, Emily S. Gurley, Stephen Luby

**Author notes:** Corresponding author (AS).

## Abstract

Antimicrobial resistance (AMR) is a growing global health threat that contributes to substantial neonatal mortality. Bangladesh has reported some of the highest rates of AMR among bacteria causing neonatal sepsis. To better understand routes of AMR transmission to newborns, we aimed to characterize the frequency of and risk factors for AMR colonization of mothers and newborns during hospitalization for delivery. We enrolled 177 pregnant women presenting for delivery to a tertiary care hospital in Faridpur, Bangladesh, during February-October 2020. We collected vaginal and rectal swabs from mothers on presentation and after delivery as well as rectal swabs from newborns. We also collected swabs from the hospital environment proximal to the patients. Swabs were plated on chromogenic agars selective for extended-spectrum-beta-lactamase producing organisms (ESBL) and carbapenem-resistant organisms (CRO). We performed univariable and multivariable analyses to determine factors associated with ESBL/CRO colonization. Prior to delivery, 17% of mothers had vaginal colonization and 71% had rectal colonization with ESBLs; 4% had vaginal colonization and 13% had rectal colonization with CROs. Seventy-nine percent of women underwent cesarean deliveries (C-section). Ninety-eight percent of women received prophylactic antibiotics during hospitalization. Following delivery, nearly 90% of mothers and newborns were colonized with ESBLs and over 70% with CROs. Of the 290 environmental samples, 77% were positive for ESBLs, and 69% were positive for CROs. Maternal and newborn colonization at discharge were both associated with C-section (RR for maternal 1.4; 95% CI 1.0-1.8 and newborn 1.3; 95% CI 1.1-1.7). Facility-based deliveries increase exposure to AMR organisms, likely driven by intense use of antibiotics and frequent C-sections. Greater attention should be given to the use of perinatal antibiotics, indications for C-sections, and infection prevention practices to reduce the high prevalence of colonization with antibiotic resistant bacteria.

## Introduction

Antimicrobial resistance (AMR) is a growing global health threat that disproportionately affects low- and middle-income countries (LMIC) and represents one of the leading causes of mortality worldwide.[1,2] Neonates are a key risk group for infections with AMR organisms given their lack of microbiome. Neonates are often first exposed to these organisms at the time of birth, which can result in colonization. Colonization occurs when bacteria persist in or on body surfaces without causing illness. Although colonization does not directly cause disease, colonization with resistant bacteria can predispose individuals to developing drug-resistant infections in the future, particularly among newborns with prematurity and low birthweight.[3– 5] Moreover, colonized individuals can still spread resistant organisms to other people and their environment. Neonatal sepsis caused by AMR organisms results in higher rates of mortality.[6] An estimated 200,000 neonatal deaths annually have been attributed to infections with AMR organisms.[1,7]

In many LMICs, the majority of cases of neonatal sepsis are caused by Gram-negative bacteria, many of which are multidrug resistant (MDR).[6,8–11] A report from 2020 demonstrated that 81% of Gram-negative bacteria causing sepsis in newborns across three neonatal care units in Bangladesh were resistant to carbapenems, one of the last line antibiotic options.[10] Similar concerning trends of increasing AMR in neonatal infections have been observed elsewhere in South and Southeast Asia, including among homebirths, demonstrating an increasing community reservoir for AMR.[12–14]

Increased availability of antibiotics has led to their overuse in human and agricultural contexts. The selective pressure created by repeated antibiotic exposure facilitates emergence and spread of resistance factors in the community.[15] Healthcare facilities, in particular, have been implicated as an important source of AMR amplification because of the associated intense antibiotic use and admixing of ill and susceptible patients.[16,17] This is further exacerbated in low-resource hospitals because of overcrowding, understaffing, inadequate hygiene and sanitation, and a lack of access to diagnostics. In these settings, antibiotics are often used liberally in healthcare facilities as a substitute for improved hygiene and sanitation, and the lack of diagnostics precludes antibiotic stewardship practices.[18,19] The convergence of abundant antibiotics and a heavily contaminated environment creates the ideal scenario for AMR organisms and genes to spread and novel mutations to arise. Understanding the environments that are promoting evolution and transmission of such organisms is an essential step towards preventing exposure, thereby avoiding downstream consequences such as resistant infections that may be difficult or impossible to treat.

The objective of this study was to estimate the burden and risk factors for colonization with extended-spectrum beta-lactamase producing (ESBL) organisms and carbapenem-resistant organisms (CRO) among mothers and newborns in the context of facility-based deliveries. As one step in the infection pathway, colonization provides a useful parameter for monitoring transmission patterns that could predispose to infection.[20–22]

## Methods

### Participant enrollment

We enrolled pregnant women presenting for delivery at a tertiary care public medical college hospital in Faridpur, Bangladesh, during February – March and August – October, 2020. A four-month interruption in enrollment (April-July) occurred as a result of data collection restrictions during the COVID-19 pandemic. A trained member of the nursing staff collected a set of vaginal and rectal swabs. The project research physician conducted interviews with the participants to obtain demographic and community exposure information about exposures that were hypothesized could be related to AMR colonization, including socioeconomic status, sanitation, animal contact, antibiotic use, and healthcare contact (Supplementary Figure 1). Over the course of the hospitalization, the research physician gathered information from the medical charts and care providers regarding treatments and interventions. Prior to hospital discharge, and at least 24 hours after delivery, the nursing staff collected a second set of vaginal and rectal swabs from the mothers and a rectal swab from their newborns. A total of 177 mother/baby pairs completed data collection (Figure 1).

**Figure 1.**
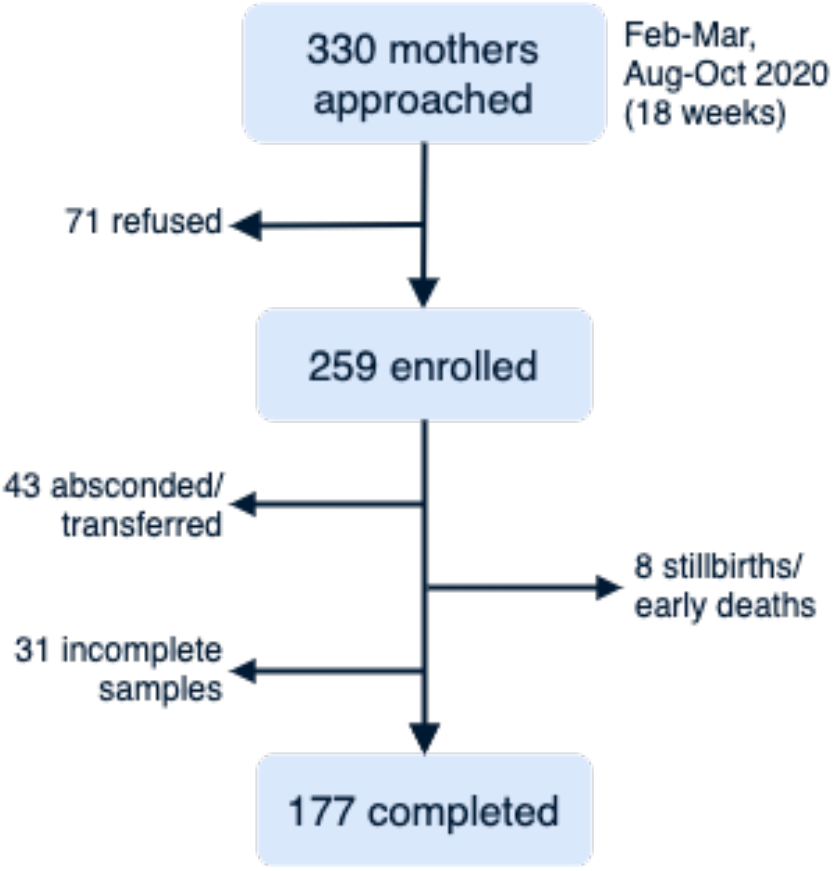
Study enrollment of pregnant mothers presenting for delivery at a tertiary care facility, Faridpur, Bangladesh, 2020. Number of pregnant women approached for enrollment, number enrolled, and number who completed all surveys and specimen collections, as well as the reasons for attrition.

### Environmental sampling

During the same time interval as participant enrollment, the study team collected samples of the hospital environment from the perinatal ward, labor room, and the operating room in which cesarean deliveries (C-section) were performed. The selection of sampled items was determined following a 3-day observation period during which frequently touched surfaces and shared equipment were identified and counts of hand contacts were noted for each type of surface or equipment. We purposefully sampled these sites throughout the study period. The sampling sites included hands of healthcare workers and patient attendants, shared medical equipment, beds, faucets, doors, toilet facilities, oxygen delivery devices, floors, and other surfaces. For each day of participant enrollment, we obtained 3 environmental swabs. Swabs were first moistened with sterile water before swabbing surfaces for a duration of 20-30 seconds.

### Bacterial culturing of swab samples

Participant (rectal and vaginal) and environmental swabs were stored at 2-8°C until they could be transported to the lab. All swabs were processed within 24 hours of collection. Participant swabs were directly inoculated onto CHROMagar™ ESBL, selective for ESBL-producing organisms, and CHROMagar mSuperCARBA (CHROMagar, Paris, France), selective for CROs. Environmental swab samples were enriched with trypticase soy broth (TSB) with overnight incubation at 37°C before the samples were inoculated onto the same media as the participant swabs. From the chromogenic agars, bacterial colony growth color and characteristics were recorded after overnight incubation at 37°C.

### Statistical analysis

We performed descriptive statistics to summarize epidemiologic characteristics of participants. We compared the proportion of colonized individuals and environmental samples before the start of the COVID-19 pandemic (Feb-Mar) and during COVID (Aug-Oct). We examined the relationship between community-based exposures on pre-delivery AMR colonization patterns and hospital-based exposures on post-delivery AMR colonization patterns (either ESBL-producing organisms, CROs, or both) using generalized linear models. We calculated unadjusted risk ratios for all community- and hospital-based exposures with at least 10% prevalence. We used McNemar’s test to determine differences in colonization prevalence across various time points for paired samples and a test of proportions to determine differences in colonization for unpaired samples.

Community or hospital exposures with p-values <0.2 that had at least 10% prevalence and were not collinear were included in multivariable analysis. No additional model refining was performed as the objective of the analysis was not to optimize a final predictive model, which would likely not be robust given the limited dataset, but to generate hypotheses regarding factors most likely to drive AMR colonization.

### Ethics

We obtained written informed consent from all adult study participants. Consent for newborn enrollment was provided by the parents and/or legal guardians. The study protocol was reviewed and approved by the IRB at Stanford University (protocol #53442) and the Research and Ethics Review Committees at icddr,b (protocol #PR-19119).

### Funding

Funding was provided by NIH FIC Global Health Equity Scholars D43 TW010540 as well as the Thrasher Research Fund (award #15188). The funders had no role in the study design, interpretation of results, or decision to publish.

## Results

### Demographics of pregnant women enrolled in the study

Of the 177 women enrolled in the study, the median age was 25 years (range 17-40) (Table 1). The majority of participants (73%) had at least a secondary school education. The median monthly household income was 201 USD (amounting to $1.34/person/day). Almost all households had improved drinking water sources, with 98% of households reporting tube wells as the main water source. Pit latrines were the most common toilet type (79%). Approximately one-third (33%) of the women had no prior pregnancies. While most women had received some prenatal care, only 14% reported 4 or more visits. Anemia was the most frequent pregnancy complication, reported by nearly half of participants. Two-thirds of deliveries occurred at or post-term.

**Table 1.**
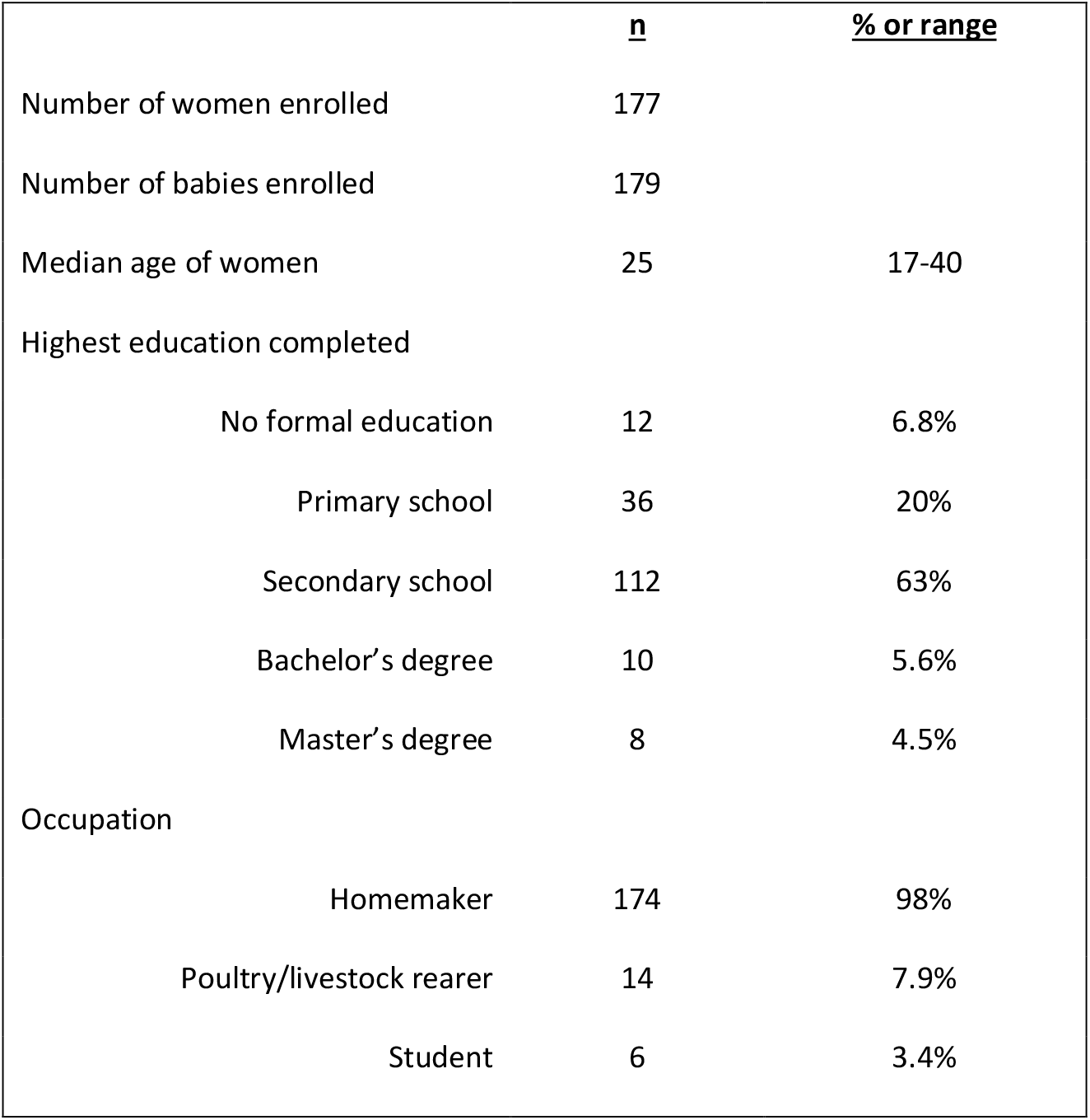
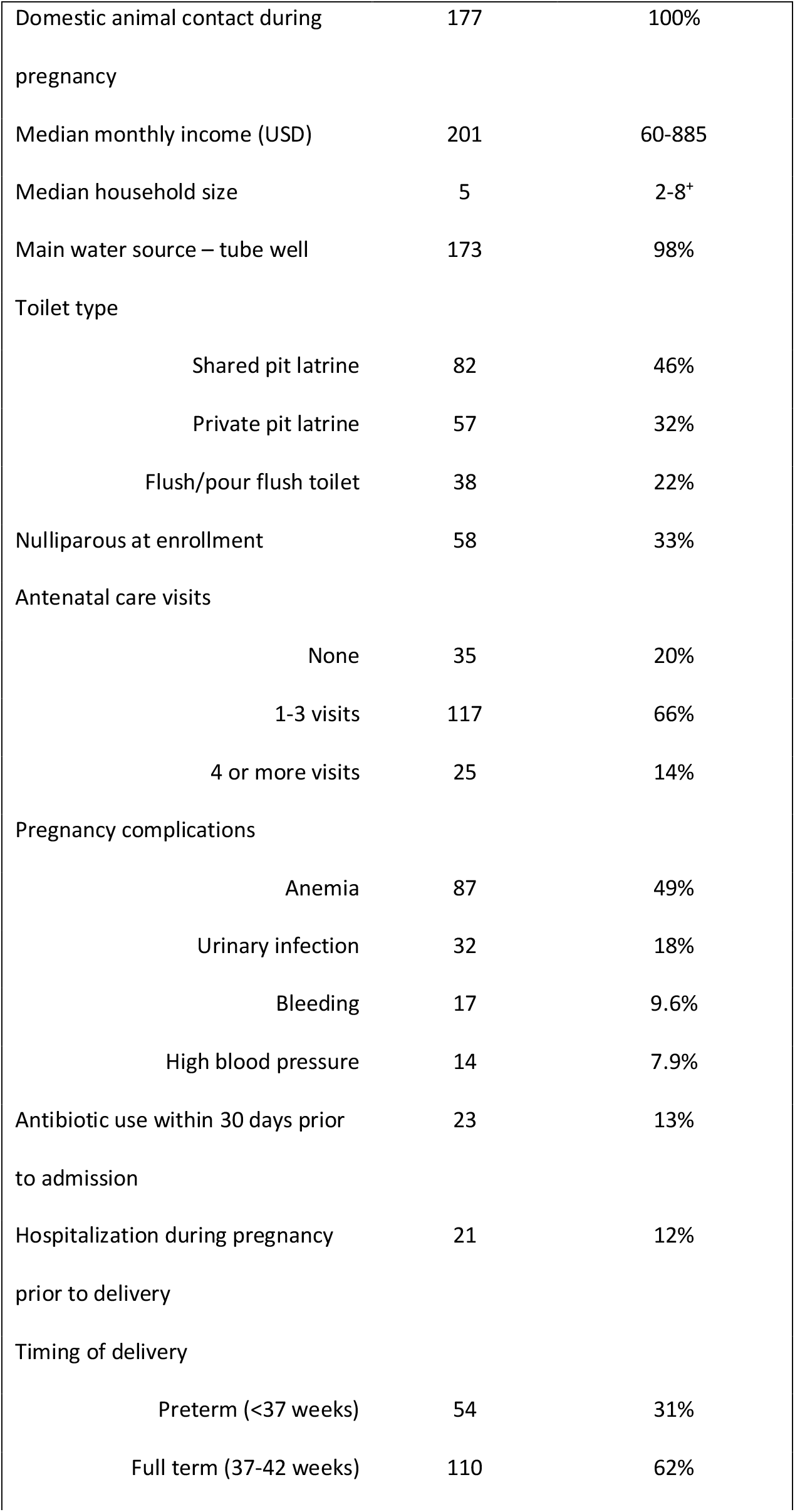
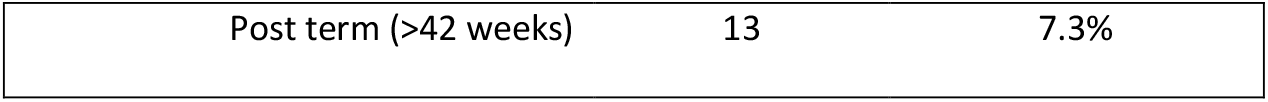
Community-based characteristics of pregnant mothers presenting for delivery, Faridpur, Bangladesh, 2020 (N=177)

|  | <u>n</u> | <u>% or range</u> |
| --- | --- | --- |
| Number of women enrolled | 177 |  |
| Number of babies enrolled | 179 |  |
| Median age of women | 25 | 17-40 |
| Highest education completed |  |  |
| No formal education | 12 | 6.8% |
| Primary school | 36 | 20% |
| Secondary school | 112 | 63% |
| Bachelor's degree | 10 | 5.6% |
| Master's degree | 8 | 4.5% |
| Occupation |  |  |
| Homemaker | 174 | 98% |
| Poultry/livestock rearer | 14 | 7.9% |
| Student | 6 | 3.4% |
| Domestic animal contact during pregnancy | 177 | 100% |
| Median monthly income (USD) | 201 | 60-885 |
| Median household size | 5 | 2-8+ |
| Main water source – tube well | 173 | 98% |
| Toilet type |  |  |
| Shared pit latrine | 82 | 46% |
| Private pit latrine | 57 | 32% |
| Flush/pour flush toilet | 38 | 22% |
| Nulliparous at enrollment | 58 | 33% |
| Antenatal care visits |  |  |
| None | 35 | 20% |
| 1-3 visits | 117 | 66% |
| 4 or more visits | 25 | 14% |
| Pregnancy complications |  |  |
| Anemia | 87 | 49% |
| Urinary infection | 32 | 18% |
| Bleeding | 17 | 9.6% |
| High blood pressure | 14 | 7.9% |
| Antibiotic use within 30 days prior to admission | 23 | 13% |
| Hospitalization during pregnancy prior to delivery | 21 | 12% |
| Timing of delivery |  |  |
| Preterm (<37 weeks) | 54 | 31% |
| Full term (37-42 weeks) | 110 | 62% |
| Post term (>42 weeks) | 13 | 7.3% |

The majority of women (79%) delivered via C-section (Table 2). Duration of hospitalization was longer for women undergoing C-section compared with vaginal delivery (4.0 versus 1.6 days, p=0.00). Nearly all women received perinatal prophylactic antibiotics (98%), of whom, 91% received them before or during delivery. The prescribed duration of antibiotics was longer for women who underwent C-section compared with vaginal delivery (10 versus 7.6 days, p=0.00). The most common antibiotics administered were metronidazole (89%), flucloxacillin (69%), and third generation cephalosporins (67%) (Supplementary Table 1). The most frequent antibiotic regimen was a 3-drug combination of a cephalosporin (any generation), metronidazole, and flucloxacillin (66%). No carbapenem use was reported. More than half of the participants experienced a pregnancy complication, including fetal distress/cord prolapse (40%), obstructed/prolonged labor (27%), or prolonged rupture of membranes (23%).

**Table 2.** Hospital-based characteristics and management of pregnant mothers presenting for delivery, Faridpur, Bangladesh, 2020 (N=177)

|  | <u>n</u> | <u>% or range</u> |
| --- | --- | --- |
| Labor management |  |  |
| Membrane sweeping/stripping | 50 | 28% |
| 6 or more vaginal exams | 45 | 25% |
| Artificial rupture of membranes | 19 | 11% |
| Mechanical cervical ripening | 14 | 7.9% |
| Mode of delivery |  |  |
| Vaginal delivery | 37 | 21% |
| Cesarean delivery | 140 | 79% |
| Mother received antibiotics | 174 | 98% |
| Duration of antibiotics, median (days)* | 10 | 3-15 |
| Timing of antibiotic initiation* |  |  |
| Before/during delivery | 158 | 91% |
| After delivery | 16 | 9.2% |
| Indication for antibiotics* |  |  |
| Prevention | 174 | 100% |
| Treatment | 1 | 0.6% |
| Delivery complications |  |  |
| Fetal distress/cord prolapse | 70 | 40% |
| Obstructed/prolonged labor | 48 | 27% |
| Prolonged rupture of membranes | 41 | 23% |
| Retained placenta | 7 | 6.4% |
| Duration of hospitalization, median (d) | 3 | 1-9 |
| Admission to ICU | 9 | 5.1% |

\*N=174, the number who received antibiotics; ICU: Intensive care unit; d: days

Among the newborns, nearly all received airway clearing, wiping and wrapping, temperature and weight measuring, and feeding initiation within the first hour of birth (Table 3). Approximately one-quarter underwent resuscitation with two newborns requiring artificial ventilation. Five percent of newborns received antibiotics, including 3 (1.7%) newborns with clinically-diagnosed neonatal sepsis. No blood cultures were collected at the time of diagnosis or during the course of treatment. No neonatal deaths occurred among enrolled newborns during the course of the study.

**Table 3.** Hospital-based characteristics and management of newborns delivered at a tertiary care facility, Faridpur, Bangladesh, 2020 (N=177*)

|  | <u>n</u> | <u>% (range)</u> |
| --- | --- | --- |
| Female | 82 | 46% |
| Birthweight, median (g) | 2900 | 1500-4200 |
| Immediate management of newborn<br>(within 1 hour of birth) |  |  |
| Cleared airway | 176 | 99% |
| Wiped and wrapped | 176 | 99% |
| Temperature and weight measured | 173 | 98% |
| Initiated feeding | 171 | 97% |
| Administered vitamin K | 98 | 55% |
| Resuscitated | 40 | 23% |
| Mechanical ventilation | 2 | 1.1% |
| Newborn received antibiotics | 9 | 5.1% |
| Duration of antibiotics, median (d)** | 3 | 1-5 |
| Indication for antibiotics** |  |  |
| Prevention | 8 | 89% |
| Treatment | 1 | 11% |
| Newborn sepsis | 3 | 1.7% |
| Duration of hospitalization, median (d) | 3 | 1-7 |
| Admission to neonatal ICU | 9 | 5.1% |

\*For the two sets of twins, one twin was randomly excluded from each set for analysis purposes.
\*\*N=9, the number of newborns who received antibiotics; d: days.

### Colonization patterns of mothers and newborns with ESBL-producing and carbapenem-resistant organisms

On admission, 17% of women (n=30) had vaginal colonization and 71% (n=125) had rectal colonization with organisms recovered from CHROMagar ESBL plates (hereafter referred to as “ESBLs”), but only 15% (n=27) had rectal or vaginal colonization with organisms recovered from CHROMagar mSuperCARBA plates (hereafter referred to as “CROs”) (Figure 2). At the time of discharge following delivery, nearly all women had either rectal or vaginal colonization with ESBLs (98%, n=174), 86% (n=153) had rectal CRO colonization, and 74% (n=130) had vaginal CRO colonization. Newborns demonstrated rectal colonization patterns similar to maternal colonization patterns on discharge: 89% (n=157) were colonized with ESBLs, and 72% (n=128) were colonized with CROs.

**Figure 2.**
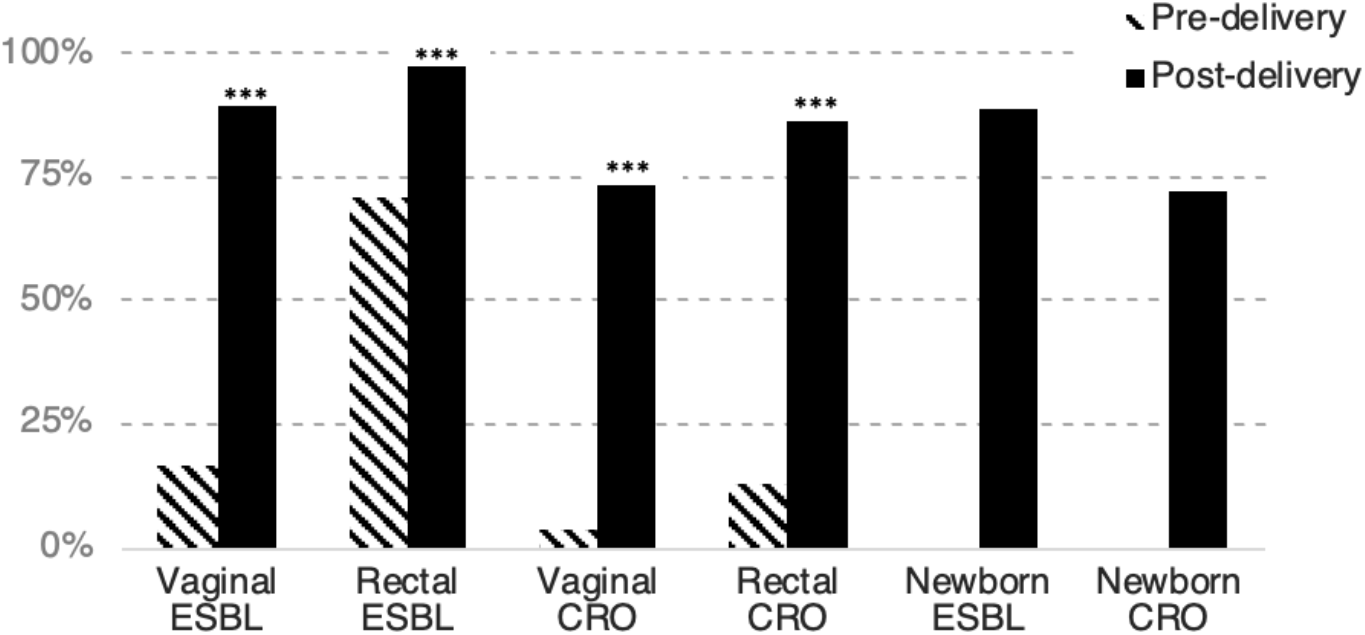
Vaginal and rectal colonization of mothers and rectal colonization of newborns with ESBL-producing and carbapenem resistant organisms at a tertiary care facility, Faridpur, Bangladesh, 2020 (N=177) Prevalence of AMR colonization patterns among mothers and newborns. Colonization was compared pre- and post-delivery to determine differences between community-based colonization and colonization following healthcare exposure. ESBL = organisms recovered from agar selective for extended-spectrum beta-lactamase-producing organisms; CRO = organisms recovered from agar selective for carbapenem resistant organisms. ***McNemar’s test p<0.001.

### Prevalence of ESBLs and CROs in environmental samples

A total of 290 environmental swab samples were collected from the perinatal ward, labor room, and operating room (Supplementary Table 2). Overall, 77% (n=222) of samples were positive for ESBLs, and 69% (n=201) were positive for CROs, and patterns were similar across types of samples analyzed (Figure 3). However, many surfaces had multiple colony morphologies on the selective plates, demonstrating broad organism diversity (Supplementary Figure 2).

**Figure 3.**
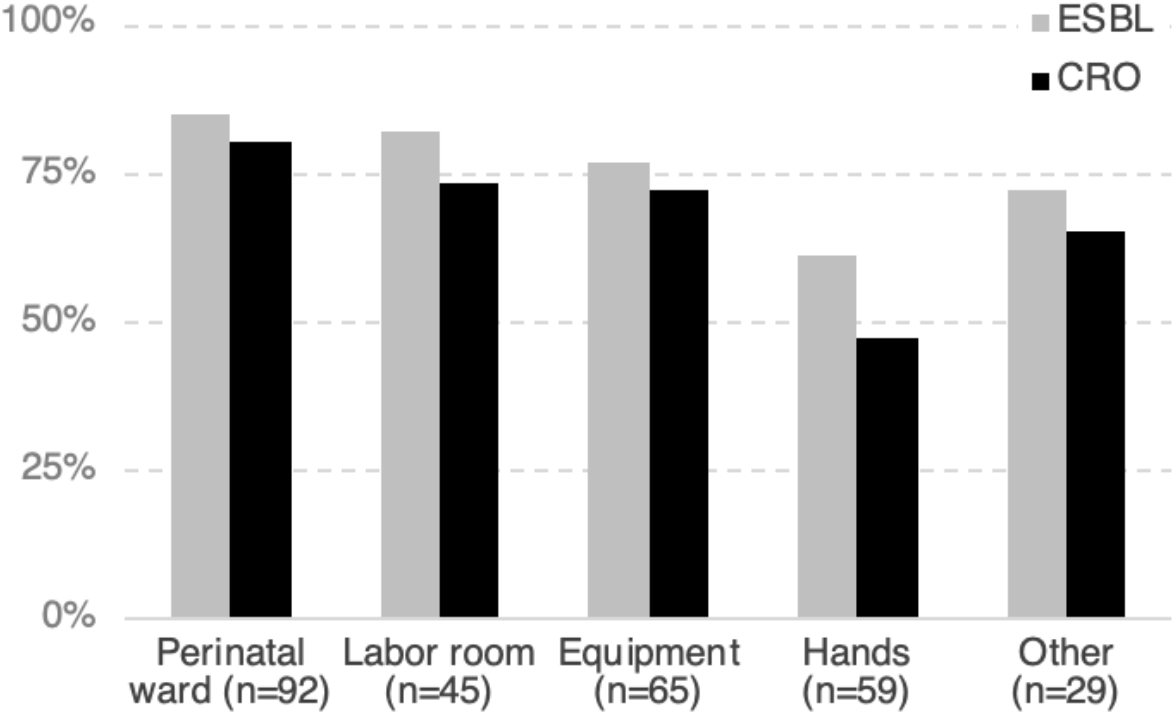
Environmental detection of ESBL-producing and carbapenem resistant organisms in the obstetric facilities of a tertiary care hospital, Faridpur, Bangladesh, 2020 (N=290) Frequency of contamination of various hospital environmental surfaces during the study period. Hand samples were taken from healthcare workers as well as patient attendants. ESBL = organisms recovered from agar selective for extended-spectrum beta-lactamase-producing organisms; CRO = organisms recovered from agar selective for carbapenem resistant organisms.

### Prevalence of ESBL/CRO colonization in relation to COVID-19

Pre-delivery rectal ESBL colonization was significantly higher after the start of the COVID-19 pandemic (80.5%) compared with before COVID (63.0%, p=0.01) (Supplementary Table 3). Similarly, rectal CRO colonization increased during COVID (8.0% vs. 19.5%, p=0.02). Pre-delivery vaginal ESBL and CRO colonization also increased, but the differences were not significant (ESBL: 15.0% vs. 19.5%, p=0.43; CRO: 2.0% vs. 6.3%, p=0.13).

In contrast, post-delivery rectal colonization was similar before and after the start of COVID (ESBL: 96.0% vs. 98.7%, p=0.28; CRO 83.0% vs. 90.9%, p=0.13). Post-delivery vaginal colonization was also stable (ESBL: 92.0% vs. 85.7%, p=0.18; CRO: 72.0% vs. 75.3%, p=0.62).

Newborn colonization increased, but the differences were only marginally significant (ESBL: 85.0% vs. 93.5%, p=0.08; CRO: 67.0% vs. 79.2%, p=0.07).

Overall environmental contamination with ESBLs and CROs was markedly higher during COVID compared with before the pandemic (ESBL: 53.9% vs. 94.4%, p=0.00; CRO: 44.5% vs. 88.9%, p=0.00) (Supplementary Figure 3).

### Community and hospital exposures associated with ESBL/CRO colonization in mothers and newborns

Colonization outcome variables that were correlated were collapsed into combined categories, except for pre-delivery rectal ESBL and rectal CRO colonization given substantial variability in prevalence (Supplementary Table 4). The pre-delivery colonization outcome variables are: vaginal ESBL and/or CRO, rectal ESBL, and rectal CRO; the post-delivery colonization outcome variables are vaginal ESBL and/or CRO, rectal ESBL and/or CRO, and newborn ESBL and/or CRO.

Hospitalization during pregnancy (apart from during delivery) was associated with an increased risk for vaginal ESBL/CRO and rectal CRO colonization across colonization categories in multivariate analyses (vaginal ESBL/CRO RR 2.1, 95% CI 1.3-3.4; rectal ESBL RR 1.3, 95% CI 1.2-1.5; rectal CRO RR 2.9, 95% CI 1.4-6.2) (Table 4). Preterm delivery was also associated with pre-delivery vaginal ESBL/CRO colonization (RR 2.6, 95% CI 1.5-4.4). Having a private pit latrine compared with a shared pit latrine was associated with less vaginal ESBL/CRO colonization (RR 0.44, 95% CI 0.24-0.81). While it did not reach statistical significance at a p<0.05 cutoff, raising ducks outside the home had the highest measures of association with pre-delivery AMR colonization (vaginal ESBL/CRO RR 2.5, 95% CI 0.78-8.0; rectal CRO RR 6.6, 95% CI 0.92-48).

**Table 4.** Association of community-based exposures with maternal pre-delivery ESBL/CRO vaginal and rectal colonization, Faridpur, Bangladesh, 2020.

|  | Vaginal ESBL/CRO |  |  |  | Rectal ESBL |  |  |  | Rectal CRO |  |  |  |
| --- | --- | --- | --- | --- | --- | --- | --- | --- | --- | --- | --- | --- |
|  | Univariable |  | Multivariable |  | Univariable |  | Multivariable |  | Univariable |  | Multivariable |  |
|  | RR | 95% CI | RR | 95% CI | RR | 95% CI | RR | 95% CI | RR | 95% CI | RR | 95% CI |
| Household size |  |  |  |  |  |  |  |  |  |  |  |  |
| <5 | Ref |  | - |  | Ref |  | - |  | Ref |  | - |  |
| 5-6 | 1 | 0.49-2.1 | - |  | 0.96 | 0.78-1.2 | - |  | 1.2 | 0.48-2.8 | - |  |
| 7 or more | 1.9 | 0.81-4.4 | - |  | 1.1 | 0.87-1.5 | - |  | 2.4 | 0.87-6.5 | - |  |
| Highest education |  |  |  |  |  |  |  |  |  |  |  |  |
| Primary or below | Ref |  | - |  | Ref |  | - |  | Ref |  | - |  |
| Secondary | 1.4 | 0.65-1.4 | - |  | 0.95 | 0.76-1.2 | - |  | 1.2 | 0.43-3.2 | - |  |
| College or above | 0.82 | 0.28-2.4 | - |  | 1 | 0.78-1.3 | - |  | 1.6 | 0.55-4.7 | - |  |
| Income |  |  |  |  |  |  |  |  |  |  |  |  |
| <12000 BDT | Ref |  | - |  | Ref |  | - |  | Ref |  | - |  |
| 12000-25000 BDT | 1 | 0.51-2.0 | - |  | 1.1 | 0.90-1.4 | - |  | 0.54 | 0.21-1.4 | - |  |
| >25000 BDT | 0.4 | 0.14-1.2 | - |  | 1 | 0.79-1.4 | - |  | 0.86 | 0.35-2.1 | - |  |
| Toilet type |  |  |  |  |  |  |  |  |  |  |  |  |
| Shared pit latrine | Ref |  | Ref. |  | Ref |  | Ref |  | Ref |  | - |  |
| Private pit latrine | 0.45 | 0.19-1.1 | **0.44** | **0.24-0.81** | 0.84 | 0.67-1.1 | 0.83 | 0.68-1.0 | 0.92 | 0.38-2.2 | - |  |
| Flush/pour flush toilet | 0.68 | 0.30-1.6 | 0.59 | 0.32-1.1 | 0.86 | 0.66-1.1 | 0.82 | 0.64-1.1 | 0.98 | 0.37-2.6 | - |  |
| Water storage |  |  |  |  |  |  |  |  |  |  |  |  |
| Open container | Ref |  | - |  | Ref |  | - |  | Ref |  | Ref |  |
| Closed container | 0.71 | 0.38-1.4 | - |  | 1 | 0.83-1.2 | - |  | 1.9 | 0.78-4.6 | 2 | 0.85-4.7 |
| Water treatment |  |  |  |  |  |  |  |  |  |  |  |  |
| Any water treatment | 0.88 | 0.42-1.9 | - |  | 0.99 | 0.80-1.2 | - |  | 1.4 | 0.61-3.0 | - |  |
| Water filter | 0.73 | 0.30-1.8 | - |  | 0.99 | 0.78-1.3 | - |  | 1.7 | 0.73-3.7 | - |  |
| Water sources |  |  |  |  |  |  |  |  |  |  |  |  |
| Rainwater | 1.4 | 0.71-2.6 | - |  | 1 | 0.83-1.3 | - |  | 0.76 | 0.32-1.8 | - |  |
| Bottled water | 1.5 | 0.65-2.4 | - |  | 1 | 0.83-1.2 | - |  | 0.83 | 0.39-1.8 | - |  |
| Goat consumption | 0.76 | 0.38-1.5 | - |  | 1.1 | 0.88-1.3 | - |  | 1.5 | 0.69-3.2 | - |  |
| Animal contact |  |  |  |  |  |  |  |  |  |  |  |  |
| Chickens inside home | 0.97 | 0.51-1.9 | - |  | 1.1 | 0.89-1.3 | - |  | 1.3 | 0.59-3.0 | - |  |
| Ducks outside home | 2.6 | 0.82-8.0 | 2.5 | 0.78-8.0 | 1.2 | 0.92-1.6 | 1.2 | 0.89-1.5 | 6 | 0.83-43 | 6.6 | 0.92-48 |
| Goats outside home | 0.76 | 0.39-1.5 | - |  | 0.92 | 0.75-1.1 | - |  | 0.65 | 0.30-1.4 | - |  |
| Cows outside home | 1.3 | 0.42-3.8 | - |  | 1.3 | 0.86-1.9 | - |  | 3 | 0.42-21 | - |  |
| Prior pregnancy | 0.77 | 0.40-1.5 | - |  | 1 | 0.82-1.2 | - |  | 0.64 | 0.29-1.4 | - |  |
| Prior miscarriage | 1.8 | 0.88-3.7 | 1.8 | 0.97-3.3 | 0.94 | 0.77-1.1 | - |  | 0.96 | 0.44-2.1 | - |  |
| Children at home |  |  |  |  |  |  |  |  |  |  |  |  |
| 0-1 | Ref |  | - |  | Ref |  | - |  | Ref |  | - |  |
| 2+ | 1.2 | 0.63-2.4 | - |  | 0.92 | 0.76-1.1 | - |  | 1.4 | 0.64-3.2 | - |  |
| Prenatal care |  |  |  |  |  |  |  |  |  |  |  |  |
| None | Ref |  | - |  | Ref |  | - |  | Ref |  | - |  |
| 1-3 visits | 0.85 | 0.39-1.9 | - |  | 1 | 0.79-1.3 | - |  | 0.65 | 0.27-1.6 | - |  |
| 4 or more visits | 0.8 | 0.26-2.5 | - |  | 0.9 | 0.62-1.3 | - |  | 0.93 | 0.29-3.0 | - |  |
| Pregnancy complications |  |  |  |  |  |  |  |  |  |  |  |  |
| Any complication | 0.75 | 0.39-1.4 | - |  | 0.95 | 0.78-1.2 | - |  | 1.3 | 0.59-3.0 | - |  |
| Anemia | 0.97 | 0.51-1.8 | - |  | 0.92 | 0.76-1.1 | - |  | 0.8 | 0.37-1.7 | - |  |
| Urinary infection | 1.09 | 0.49-2.4 | - |  | 0.97 | 0.75-1.3 | - |  | 1.3 | 0.51-3.1 | - |  |
| Hospitalization during pregnancy | **2.2** | **1.1-4.4** | **2.1** | **1.3-3.4** | 1.2 | 0.93-1.5 | **1.3** | **1.2-1.5** | **3.3** | **1.5-7.0** | **2.9** | **1.4-6.2** |
| Antibiotic use within 30 days | 0.99 | 0.38-2.6 | - |  | 1.1 | 0.89-1.4 | - |  | 1.4 | 0.5-3.8 | - |  |
| Preterm delivery | **2.1** | **1.1-4.0** | **2.6** | **1.5-4.4** | 1.1 | 0.92-1.4 | - |  | 1.5 | 0.67-3.2 | - |  |
| Vaginal ESBL/CRO | N/A |  | N/A |  | 0.9 | 0.68-1.3 | - |  | 1.7 | 0.71-3.9 | - |  |
| Rectal ESBL | 0.76 | 0.39-1.5 | - |  | N/A |  | N/A |  | ND |  | - |  |
| Rectal CRO | 1.6 | 0.74-3.5 | - |  | ND |  | - |  | N/A |  | N/A |  |

Includes all variables with at least 10% prevalence. Multivariable analysis includes all non-collinear variables with $p < 0.2$ . Results in bold have $p < 0.05$ level. RR = relative risk; ESBL = organisms recovered from agar selective for extended-spectrum beta-lactamase-producing organisms; CRO = organisms recovered from agar selective for carbapenem resistant organisms. N/A = not applicable; ND = not defined.

Among hospital exposures, vaginal ESBL/CRO colonization and newborn ESBL/CRO colonization were associated with C-section (vaginal ESBL/CRO RR 1.4, 95% CI 1.0-1.8; newborn ESBL/CRO RR 1.3, 95% CI 1.1-1.7) (Table 5). Because duration of hospitalization was collinear with type of delivery, it was not included in the multivariable analysis. Similarly antibiotics administered to mothers before delivery was also collinear with type of delivery and was thus excluded from multivariable analysis. Newborn ESBL/CRO colonization was not associated with maternal post-delivery colonization, though newborn colonization was associated with pre-delivery rectal CRO colonization (RR 1.1, 95% CI 1.1-1.2). No other hospital-based exposures were found to be significantly associated with AMR colonization.

**Table 5.** Association of hospital-based exposures with maternal and newborn ESBL/CRO vaginal and rectal colonization, Faridpur, Bangladesh, 2020.

|  | Vaginal ESBL/CRO |  |  |  | Rectal ESBL/CRO |  |  |  | Newborn ESBL/CRO |  |  |  |
| --- | --- | --- | --- | --- | --- | --- | --- | --- | --- | --- | --- | --- |
|  | Univariable |  | Multivariable |  | Univariable |  | Multivariable |  | Univariable |  | Multivariable |  |
|  | RR | 95% CI | RR | 95% CI | RR | 95% CI | RR | 95% CI | RR | 95% CI | RR | 95% CI |
| Delivery mode |  |  |  |  |  |  |  |  |  |  |  |  |
| Vaginal delivery | Ref |  | Ref |  | Ref |  | Ref |  | Ref |  | Ref |  |
| Cesarean delivery | **1.4** | **1.1-1.8** | **1.4** | **1.0-1.8** | 1.1 | 0.97-1.2 | 0.99 | 0.97-1.0 | **1.3** | **1.1-1.7** | **1.3** | **1.1-1.7** |
| Duration of hospitalization |  |  |  |  |  |  |  |  |  |  |  |  |
| <3 days | Ref |  | - |  | Ref |  | - |  | Ref |  | - |  |
| 3-4 days | **1.4** | **1.1-1.9** | - |  | 1.1 | 0.98-1.2 | - |  | **1.3** | **1.1-1.7** | - |  |
| 5 or more days | **1** | **1.0-1.7** | - |  | 1.1 | 0.95-1.2 | - |  | 1.3 | 0.98-1.7 | - |  |
| Birth management |  |  |  |  |  |  |  |  |  |  |  |  |
| None | **1.2** | **1.0-1.4** | - |  | ND |  | - |  | N/A |  | N/A |  |
| Membrane stripping/sweeping | **0.8** | **0.68-0.95** | - |  | 0.96 | 0.89-1.0 | - |  | N/A |  | N/A |  |
| 6 or more vaginal exams | **0.83** | **0.71-0.98** | - |  | 0.98 | 0.91-1.1 | - |  | N/A |  | N/A |  |
| Artificial rupture of membranes | 0.87 | 0.69-1.11 | - |  | 0.98 | 0.87-1.1 | - |  | N/A |  | N/A |  |
| Maternal complications |  |  |  |  |  |  |  |  |  |  |  |  |
| Any complications | 1.1 | 0.90-1.4 | - |  | 1 | 0.92-1.1 | - |  | N/A |  | N/A |  |
| Premature rupture of membranes | 0.93 | 0.81-1.1 | - |  | 0.97 | 0.89-1.0 | - |  | N/A |  | N/A |  |
| Obstructed labor | 1 | 0.92-1.1 | - |  | 1.0 | 0.96-1.1 | - |  | N/A |  | N/A |  |
| Fetal distress | 1.1 | 0.98-1.2 | 0.99 | 0.93-1.1 | 0.99 | 0.94-1.0 | - |  | N/A |  | N/A |  |
| Timing of maternal complications |  |  |  |  |  |  |  |  |  |  |  |  |
| Before delivery | 1.1 | 0.94-1.4 | - |  | 1.1 | 0.95-1.2 | - |  | N/A |  | N/A |  |
| At time of delivery | 0.93 | 0.85-1.0 | 1.0 | 0.96-1.1 | 0.98 | 0.93-1.0 | - |  | N/A |  | N/A |  |
| Antibiotics administered to mother before delivery | 1.2 | 0.94-1.6 | - |  | ND |  | - |  | 1.2 | 0.93-1.6 | - |  |
| Mother received 3rd generation cephalosporin | 1.1 | 0.98-1.3 | 1.1 | 0.95-1.2 | 1.0 | 0.96-1.1 | - |  | 1.1 | 0.94-1.2 | - |  |
| Low birthweight | N/A |  | N/A |  | N/A |  | N/A |  | 0.97 | 0.83-1.1 | - |  |
| Postnatal management of newborn |  |  |  |  |  |  |  |  |  |  |  |  |
| Skin-to-skin or kangaroo care | N/A |  | N/A |  | N/A |  | N/A |  | 0.95 | 0.85-1.1 | - |  |
| Administration of vitamin K | N/A |  | N/A |  | N/A |  | N/A |  | 0.98 | 0.88-1.1 | - |  |
| Resuscitation | N/A |  | N/A |  | N/A |  | N/A |  | 0.95 | 0.82-1.1 | - |  |
| Pre-delivery vaginal ESBL/CRO colonization | 1.0 | 0.89-1.2 | - |  | 0.99 | 0.93-1.1 | - |  | 0.93 | 0.79-1.1 | - |  |
| Pre-delivery rectal ESBL colonization | 1.0 | 0.92-1.2 | - |  | 1.0 | 0.97-1.1 | - |  | 0.97 | 0.87-1.1 | - |  |
| Pre-delivery rectal CRE colonization | 0.92 | 0.75-1.1 | - |  | ND |  | - |  | 1.1 | 0.98-1.2 | **1.1** | **1.1-1.2** |
| Post-delivery vaginal ESBL/CRO colonization | N/A |  | N/A |  | 1.2 | 0.96-1.4 | 1.2 | 0.97-1.5 | 1.2 | 0.93-1.6 | - |  |
| Post-delivery rectal ESBL/CRO colonization | 2.3 | 0.77-6.7 | 1.4 | 0.76-2.6 | N/A |  | N/A |  | 1.5 | 0.73-1.1 | - |  |

Includes all variables with at least 10% prevalence. Multivariable analysis includes all non-collinear variables with $p < 0.2$ . Results in bold have $p < 0.05$ level. RR = relative risk; ESBL = organisms recovered from agar selective for extended-spectrum beta-lactamase-producing organisms; CRO = organisms recovered from agar selective for carbapenem resistant organisms. N/A = not applicable; ND = not defined.

## Discussion

This study revealed a high prevalence of perinatal colonization with ESBLs and CROs among mothers and newborns undergoing facility-based deliveries at a tertiary care hospital in Bangladesh. Colonization prevalence was substantially higher in mothers at the time of discharge compared with admission, particularly for CROs, perhaps because ESBL colonization was already frequent on admission. Additionally, there were concurrently high rates of C-sections and prescribing of prolonged courses of prophylactic antibiotics.

Newborn colonization prevalence more closely resembled maternal colonization at discharge compared with admission and occurred within the context of widespread environmental contamination. A similarly high prevalence of ESBL colonization in newborns has been reported from studies in India, Cambodia, Madagascar, and Tanzania, though CRO colonization was infrequent.[23–26] Another study from Bangladesh revealed that 82% of healthy infants were colonized with *Escherichia coli* resistant to third-generation cephalosporins.[27] Much lower neonatal AMR colonization rates have been reported in high income countries such as Israel and Sweden where ESBL colonization ranged from 5-14%.[28,29] This may reflect differences in local epidemiology as well as facility-based practices. This study has revealed one of the highest reported prevalences of CRO colonization among newborns, which is particularly concerning given the lack of effective treatment options for infections with these organisms. It is likely these organisms are leading to infections given studies showing high levels of CROs causing neonatal sepsis in Bangladesh.[10] Furthermore, the remarkably high prevalence of CRO colonization occurred despite no reported use of carbapenem antibiotics, demonstrating that other beta-lactam antibiotics may be promoting CRO colonization.

This particular hospital is a tertiary care facility where many high-risk pregnancies are referred, which may partially explain the high rates of C-sections. However, this also reflects national trends in Bangladesh. In 2018, one-third of deliveries occurred by C-section, including 67% of facility-based deliveries.[30] This is well above the 10-15% recommendation by WHO for rates of C-section.[31] The WHO threshold is based on a review of data demonstrating no improvement in perinatal mortality for higher rates of C-section.[32] This trend of increasing C-sections is growing fastest in LMICs.[33] Rather than reflecting improved global access to a life-saving procedure, the disparities in rates of C-sections - from 5% in sub-Saharan Africa to 43% in Latin America and the Caribbean - reveals ongoing incongruencies and inappropriate surgical stewardship. By 2030, nearly a third of all deliveries globally are expected to occur by C-section, with the vast majority (nearly 90%) occurring in LMICs.

Along with frequent C-sections, perinatal antibiotic use was high among participants. Despite WHO guidance recommending only a single dose of pre-operative prophylactic antibiotics for C-sections, most participants who underwent C-section were advised to complete prolonged courses of antibiotics.[34] Moreover, the WHO guidance emphasizes the use of narrow-spectrum antibiotics for prophylaxis. Reports from India demonstrate prolonged courses of prophylactic antibiotics being given to 80% women undergoing C-section, including frequent use of 3-drug regimens.[35] A study from China revealed that 100% of women who underwent C-section received a standard 7-day antibiotic regimen.[36] Similar practices have been reported from other LMICs, indicating the use of prolonged prophylactic antibiotics for C-section may be widespread, despite evidence showing no benefit from multiple doses of antibiotics.[37–40] This contrasts with practices in high-resource contexts, such as the U.S., where multiple doses of antibiotics is rare.[41,42]

Although the analysis conducted here was exploratory, prior hospitalization was consistently associated with increased risk for maternal ESBL/CRO colonization, implicating the role of healthcare settings in the propagation of AMR. Additionally, there was an association between preterm delivery and maternal vaginal ESBL/CRO colonization but not rectal colonization. This suggests that the vaginal microbiome may be having an effect on preterm delivery, which has been previously demonstrated.[43,44] It remains unclear what the mechanism is for how colonization with resistant organisms modulates preterm delivery. However, it has been proposed that antibiotic resistant bacteria may be associated with more inflammation and perhaps reflective of dysbiosis.[45,46]

The protective effects associated with a private pit latrine (as opposed to a shared pit latrine) on maternal ESBL/CRO colonization points to a hygiene component to community-based colonization. This is consistent with prior studies demonstrating that shared sanitation carries increased risk of exposure to pathogenic organisms, particularly AMR organisms.[47,48] However, use of a shared pit latrine could also be a marker for low socioeconomic status, and there could be other elements associated with socioeconomic status that are driving this association. Previous studies from South Asia have shown that low socioeconomic status is associated with increased risk for bacteriuria with antibiotic resistant bacteria in pregnancy.[49]

Duck rearing was associated with increased risk across colonization patterns. Though this association was not statistically significant at p<0.05, the relatively low number of participants with this exposure may have contributed to this, and the pattern remains intriguing. No other animal exposures carried such an association, mainly because there was little diversity among other animal exposure types. Ducks, unlike other animals raised for food, have wider geographic ranges that include surface water bodies. Notably, other studies from Bangladesh have found high prevalence of ESBL *E. coli* in rural pond and river waters.[50] This same study observed higher prevalence of ESBL *E. coli* in mixed poultry fecal litter, which included droppings from chickens as well as ducks, compared with chicken fecal litter alone.

Furthermore, ducks have more liquid feces and higher volume of feces compared to chickens, potentially resulting in greater environmental spread. This finding may warrant further investigation into the One Health processes that could be contributing to AMR propagation and dissemination.

C-section was the only hospital-based exposure associated with post-delivery maternal ESBL/CRO colonization in the multivariable analysis, though prolonged hospital stay was also significant in univariable analysis. Because these factors were collinear, length of hospital stay was not included in the multivariable analysis. These same trends were observed for newborn colonization. This is consistent with another study in a low-resource context examining associations with neonatal AMR colonization that found aspects of the healthcare setting to be the most highly associated factors, including antibiotic use, longer hospital stays, prematurity, and lower staffing ratios.[51] Additionally, C-sections have been shown to lead to a disrupted colonizing microbiota among neonates,[52–54] which may be exacerbated by antibiotic use.[55] Even when the antibiotics are only administered to the mother, studies have demonstrated negative health outcomes in the newborn, including necrotizing enterocolitis,[56] neonatal sepsis,[57] and growth stunting.[55] Further, the magnitude of disruption on the neonatal microbiome attributable to intrapartum antibiotics administered to mothers has been found to be similar to postnatal antibiotics administered to newborns directly.[58] Thus, ESBL and/or CRO colonization may be a marker of dysbiosis, induced by upstream factors such as antibiotic exposure that provide an opportunity for colonization with resistant bacteria.[46,59] Fecal microbiota transplantation has been tried as a successful strategy for restoring the microbiota in neonates born by C-section.[60] In this study, the impact of maternal antibiotic use on newborn colonization could not be assessed since nearly all women received perinatal antibiotics.

While there are likely many aspects of the healthcare setting contributing to ESBL/CRO colonization, both antibiotic use and C-section rates are potentially modifiable. Apart from contributing to AMR, unnecessary C-sections may have additional negative health ramifications, such as surgical site infections, blood clots, or injury to other organs. A review of indications for C-sections in Bangladesh found that the majority were not performed out of medical necessity.[61] This is a clear area for ongoing surveillance and changes to current practices.

Overuse of antibiotics is a known driver of AMR. The prolonged antibiotic courses received by the majority of mothers for the purposes of prophylaxis pose a threat to community rates of AMR through direct and indirect effects on resistance.[62] However, the situation in the delivery ward is unlikely to be unique in Bangladesh given high levels of antibiotic resistance reported across a variety of settings.[63,64] Reasons for liberal antibiotic use in Bangladesh include lack of understanding of antibiotic function, low awareness of antibiotic resistance, overemphasis on use of antibiotics for prevention, and a perception of antibiotics as a symbol of power.[65] Additionally, this tertiary care facility does not have a functioning microbiology laboratory or infectious disease consultant, limiting the ability to select appropriate antibiotics based on antibiograms or culture results or seek expert consultation.

The high level of antibiotic use for prophylaxis also points to a heightened concern for healthcare-associated infections (HAIs). Hospital-acquired neonatal infections are up to 20 times more common in LMICs compared with high-income contexts.[66] Accordingly, Zaidi, et al note in the context of LMICs, any newborn infection in a hospital-born baby should be considered a HAI, regardless of the timing of onset.[66] In LMICs such as Bangladesh, antibiotics are a readily-available tool to mitigate HAIs, certainly more accessible than infrastructure changes to improve hygiene and sanitation or even incentives for hospital workers to prioritize cleanliness. In this scenario, antibiotics are likely acting both as a substitute for and an extension of infection prevention and control efforts.[19] In fact, a prior study from Bangladesh demonstrated that antibiotics given to patients on admission resulted in less hospital-onset diarrhea resulting from contaminated food.[67]

Simultaneous recovery of abundant ESBLs/CROs from the hospital environment further suggests that the environment is likely contributing to AMR transmission. This has been corroborated by other studies in similar contexts that have shown that surface colonization predicts infecting organisms in neonates.[68,69]. Antibiotic resistant *Klebsiella pneumoniae* strains isolated from newborns hospitalized in a neonatal intensive care unit were found to be genetically closely related to strains isolated from other infants on the ward, supporting the notion of transmission being driven by healthcare workers and/or shared equipment.[51] Another study found that while *K. pneumoniae* transmission to newborns appears to be driven by direct contact with colonized healthcare workers, spread of *Escherichia coli* and *Enterobacter cloacae* are mediated through indirect contamination.[24] Further supporting the linkage between exposure to the healthcare environment and ESBL/CRO colonization is the increased environmental contamination during COVID-19 and concomitant increase in neonatal AMR colonization. Increased environmental contamination during COVID-19 may have resulted from decreased routine cleaning by staff who were concerned about heightened exposure to SARS-CoV-2 in the hospital setting. This supports the need for enhanced environmental cleaning and infection prevention efforts to reduce HAIs and AMR.

Some of the strengths of this study are that it employs a relatively low-cost method of AMR surveillance that could be replicated in other low-resource contexts. It also uses colonization to detect AMR burden and transmission, allowing for preventive measures to be implemented in advance of infectious outcomes. Further, the examination of both community and hospital-related factors and the measurement of colonization at two time points allows for better triangulation of the most important risk factors for AMR colonization in both settings.

This study has several limitations. The homogeneity of participants’ community and hospital exposures that we hypothesized to have strong associations with AMR colonization meant that the associations between some factors and ESBL/CRO colonization could not be evaluated.

However, the descriptive characteristics provide insight on common practices that may be implicated in AMR transmission, such as abundant antibiotic use.

Additionally, we only measured colonization status, which does not in itself carry negative health consequences and may rapidly change after returning to the home environment. Yet, other studies have shown that neonatal colonization patterns can persist for up to 5 years, with greater persistence of more virulent bacterial strains.[28,70] A meta-analysis examining the relationship between Gram-negative bacterial colonization and bloodstream infections in neonates did not find a statistically significant correlation, though the analysis was limited by a small number of studies to draw from and high heterogeneity between studies.[4] However, subsequent studies and other studies not included in the meta-analysis have supported the role of intestinal colonization as a predisposing factor to infection.[23,26,68,71]

In this study, we identified ESBL/CRO colonization by using direct culture of rectal/vaginal swabs on chromogenic agars, which is an imperfect method of classification. However, we did not use the chromogenic findings as a means of definitive identification, and instead highlighted the relative changes between pre-and post-delivery as comparable and informative indicators of underlying trends. Our findings do not prove that the environment was the source of ESBL/CRO colonization among mothers and neonates as we lack bacterial characterization data. Regardless, the remarkable abundance of ESBLs/CROs throughout the hospital environment suggests this is likely to be a factor in at least some of the transmission pathways. Future studies including whole genome sequencing-based characterization of isolates would add further clarity to transmission dynamics and AMR diversity in this setting.

## Conclusion

The scenario of rising AMR among newborns is becoming increasingly common across LMICs and demands a close examination of the factors surrounding hospital-based deliveries, including indications for C-sections and antibiotic administration. The findings of this study highlight the need to avoid overmedicalization of deliveries. Pregnant women presenting to a hospital for delivery may be more likely to be construed as “patients in need of treatment”. Separating routine perinatal care from a healthcare facility to an adjacent birthing center may help decrease the treatment imperative. Conducting randomized controlled trials demonstrating the non-inferiority of reduced antibiotic use could further support changes in antibiotic prescribing. However, this may not be supported without concomitant improvements in infection prevention and control coupled with enhanced access to diagnostics and microbiologic laboratory capacity. Unnecessary C-sections must also be curtailed to reduce the disruption of the newborn microflora and associated morbidity. Interventions to reduce C-sections could focus on understanding and dismantling the incentives driving increasing rates of C-sections. Overall, these findings demonstrate the urgent and pressing need for better AMR surveillance and associated interventions to ensure safer birthing environments.

## Data Availability

All relevant data are within the manuscript and its Supporting Information files.

## Supplementary Materials

**Supplementary Table 1.** Maternal antibiotics and prophylactic antibiotic regimens (N=174)

| **Antibiotic** | **N (%)** |
| --- | --- |
| Flucloxacillin | 120 (69.0%) |
| 1<sup>st</sup> gen cephalosporin | 25 (14.3%) |
| 2<sup>nd</sup> gen cephalosporin | 33 (18.9%) |
| 3<sup>rd</sup> gen cephalosporin | 116 (66.6%) |
| Cephalosporin/beta-lactamase inhibitor | 2 (1.1%) |
| Ciprofloxacin | 1 (0.6%) |
| Aminoglycoside | 2 (1.1%) |
| Metronidazole | 154 (88.5%) |
| **Antibiotic Regimens** |  |
| 3<sup>rd</sup> gen cephalosporin + metronidazole + flucloxacillin | 84 (48.3%) |
| 2<sup>nd</sup> gen cephalosporin + metronidazole + flucloxacillin | 33 (19.0%) |
| 1<sup>st</sup> gen cephalosporin + metronidazole | 25 (14.4%) |

**Supplementary Table 2.** Surfaces sampled for environmental swab collection.

| **Category** | **% Positive for ESBL** | **% Positive for CRO** | **Notes** |
| --- | --- | --- | --- |
| Ward (n=36) | 0.9 | 0.8 | Nurses' desk, doctors' table, doors, sink faucet, patient bench, patient chair |
| Labor room (n=31) | 0.8 | 0.6 | Labor bed cover, delivery apron, delivery shoes, delivery room light, sink faucet, sink basin, doors, chair |
| OR (n=9) | 0.6 | 0.4 | Operating room table, light handle, OR shoes, IV pole, oxygen mask, gurney |
| Newborn surfaces (n=37) | 0.8 | 0.8 | Newborn scale, newborn resuscitation tray, newborn oxygen mask, newborn suction device |
| HCW hands (n=48) | 0.5 | 0.4 | Doctors, nurses, midwives, trainees/students |
| Bed (n=26) | 0.9 | 0.8 | Bed rails, mattresses, linens |
| Floor (n=29) | 0.9 | 0.9 | Perinatal ward, labor room, stairs, dressing room |
| Attendant hands (n=11) | 0.9 | 0.7 | Patient attendants' hands |
| Toilet (n=16) | 0.9 | 0.9 | Water tap, bodna (cleaning vessel), door |
| Gurney (n=7) | 1.0 | 1.0 | Gurneys for patient transport |
| Medicine (n=12) | 0.8 | 0.8 | Medicine bottles, medicine delivery tray |
| Equipment (n=19) | 0.6 | 0.5 | Stethoscope, IV pole, scissors, equipment tray |
| Other (n=9) | 0.7 | 0.4 | Cat (lives in perinatal ward), patient mobile phone, money, patient food table, food delivery cart |

ESBL = organisms recovered from agar selective for extended-spectrum beta-lactamase-producing organisms; CRO = organisms recovered from agar selective for carbapenem resistant organisms.

**Supplementary Table 3.** Prevalence of ESBL/CRO colonization before and during COVID-19.

|  | <u>Before COVID</u> | <u>During COVID</u> | <u>p-value</u> |
| --- | --- | --- | --- |
| Pre-vaginal ESBL | 15.0% | 19.4% | 0.43 |
| Pre-rectal ESBL | **63.0%** | **80.5%** | **0.01** |
| Pre-vaginal CRO | 2.0% | 6.5% | 0.13 |
| Pre-rectal CRO | **8.0%** | **19.5%** | **0.02** |
| Post-vaginal ESBL | 92.0% | 85.7% | 0.18 |
| Post-rectal ESBL | 96.0% | 98.7% | 0.28 |
| Post-vaginal CRO | 72.0% | 75.3% | 0.62 |
| Post-rectal CRO | 83.0% | 90.9% | 0.13 |
| Newborn ESBL | 85.0% | 93.5% | 0.08 |
| Newborn CRO | 67.0% | 79.2% | 0.07 |

Results in bold are significant at the $p < 0.05$ level. ESBL = organisms recovered from agar selective for extended-spectrum beta-lactamase-producing organisms; CRO = organisms recovered from agar selective for carbapenem resistant organisms.

**Supplementary Table 4.** Correlations of outcome variables.

|  | Prevag<br>ESBL | Prerectal<br>ESBL | Prevag<br>CRO | Prerectal<br>CRO | Postvag<br>ESBL | Postvag<br>CRO | Postrectal<br>ESBL | Postrectal<br>CRO | Newborn<br>ESBL | Newborn<br>CRO |
| --- | --- | --- | --- | --- | --- | --- | --- | --- | --- | --- |
| Prevag ESBL | 1 |  |  |  |  |  |  |  |  |  |
| Prerectal<br>ESBL | -0.0723 | 1 |  |  |  |  |  |  |  |  |
| Prevag CRO | 0.3719 | 0.0672 | 1 |  |  |  |  |  |  |  |
| Prerectal<br>CRO | 0.0941 | 0.2493 | 0.1802 | 1 |  |  |  |  |  |  |
| Postvag<br>ESBL | 0.0594 | 0.0568 | -0.0233 | -0.0831 | 1 |  |  |  |  |  |
| Postvag<br>CRO | -0.0353 | 0.1177 | 0.0564 | 0.0041 | 0.5767 | 1 |  |  |  |  |
| Postrectal<br>ESBL | -0.0139 | 0.1146 | 0.0346 | 0.0659 | 0.2714 | 0.2063 | 1 |  |  |  |
| Postrectal<br>CRO | -0.085 | 0.0344 | 0.0804 | 0.0549 | 0.1825 | 0.4719 | 0.4305 | 1 |  |  |
| Newborn<br>ESBL | -0.0766 | -0.0343 | 0.0724 | 0.0849 | 0.1645 | 0.1895 | 0.1546 | 0.1714 | 1 |  |
| Newborn<br>CRO | -0.158 | 0.0999 | -0.004 | 0.0889 | 0.1934 | 0.2856 | 0.1231 | 0.3082 | 0.5769 | 1 |

ESBL = organisms recovered from agar selective for extended-spectrum beta-lactamase-producing organisms; CRO = organisms recovered from agar selective for carbapenem resistant organisms.

**Supplementary Figure 1.**
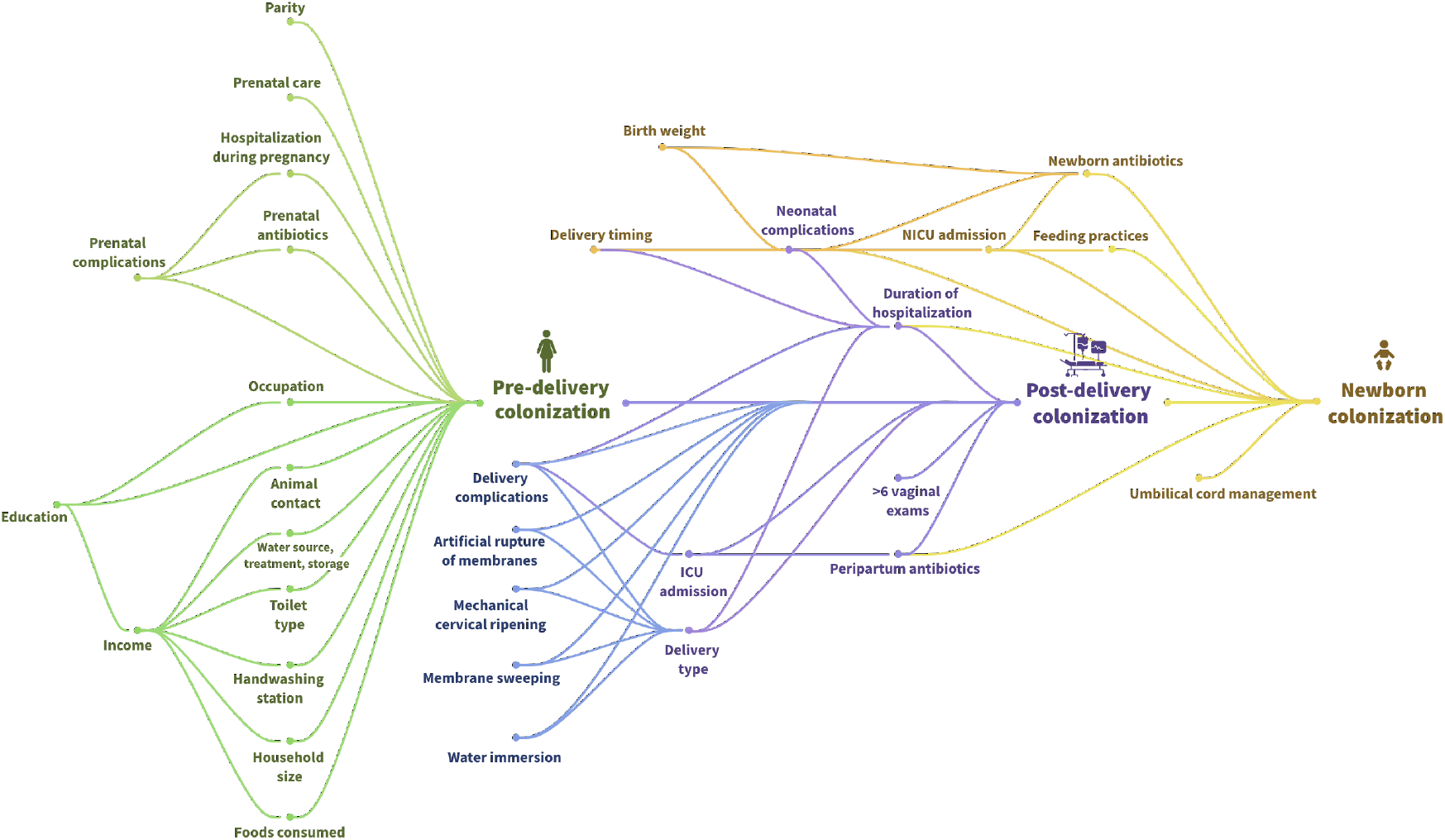
Causal pathway. A proposed causal pathway examining the relationship between community and hospital-based factors and colonization with antimicrobial resistant organisms.

**Supplementary Figure 2.**
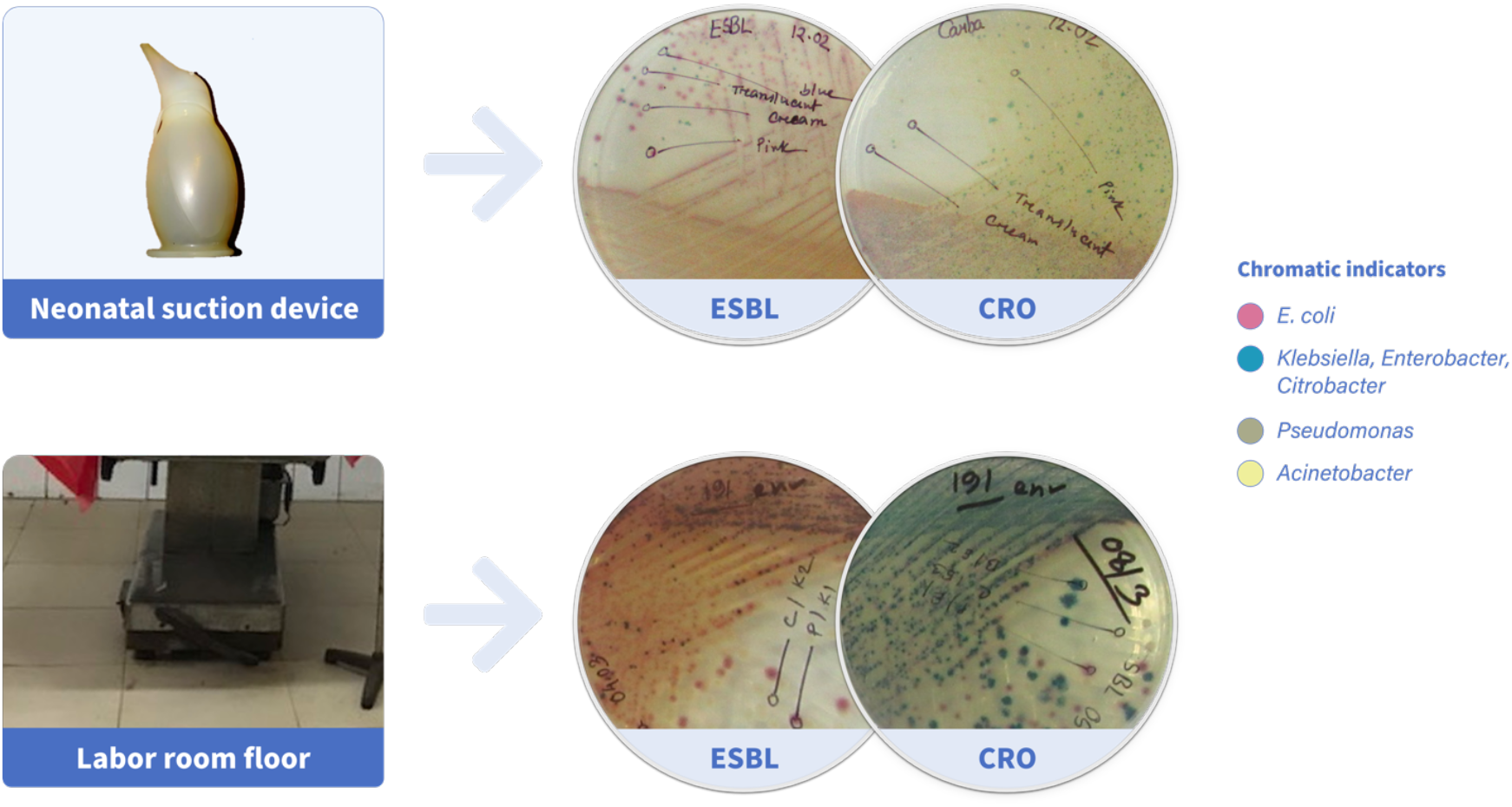
Bacterial growth from different environmental surfaces. The above panels show the bacteriologic results from some of the environmental samples. The top panel demonstrates 4 separate ESBL colony types and 3 CRO colony types. The bottom panel reveals a similar wide diversity of AMR organisms isolated from the labor room floor. ESBL = organisms recovered from agar selective for extended-spectrum beta-lactamase-producing organisms; CRO = organisms recovered from agar selective for carbapenem resistant organisms.

**Supplementary Figure 3.**
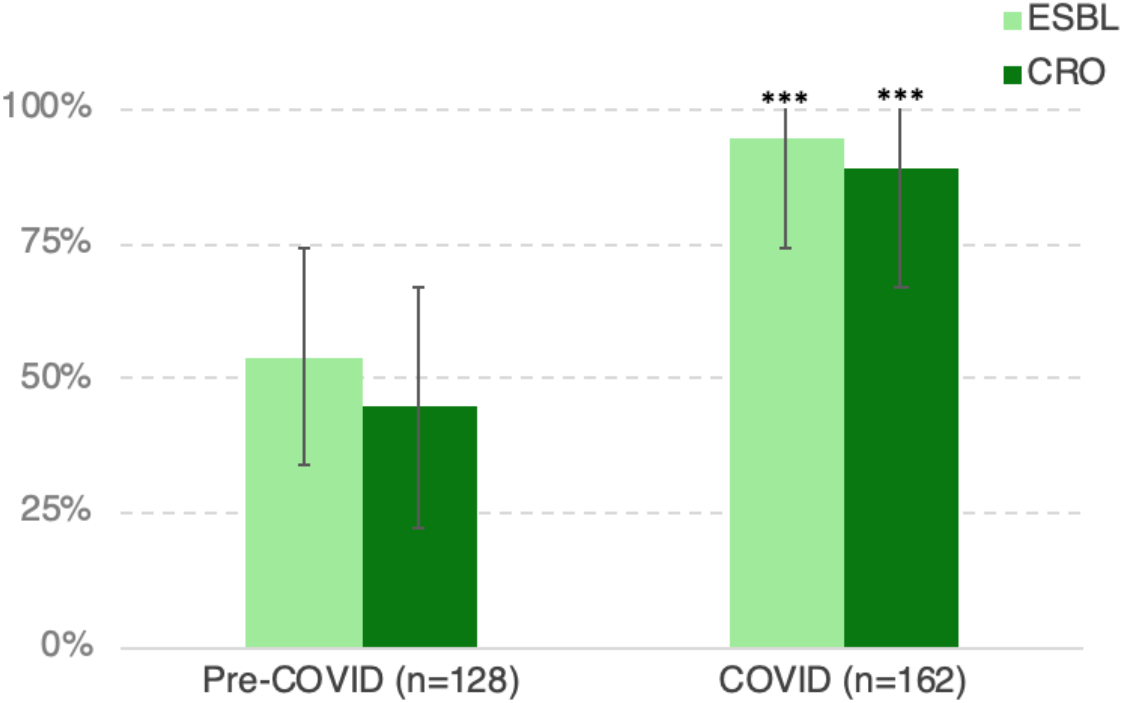
Environmental contamination with ESBLs/CROs before and during COVID-19. Environmental sampling demonstrated increased contamination with antimicrobial resistant organisms after the onset of the COVID-19 pandemic compared with before the pandemic. ESBL = organisms recovered from agar selective for extended-spectrum beta-lactamase-producing organisms; CRO = organisms recovered from agar selective for carbapenem resistant organisms. *\*\*\*significant at the p<0*.*0001 level*

